## Supplementary material for "Development of a vocal biomarker for fatigue monitoring in people with COVID-19": SOM 1. Text to read

**Supplementary Online Material 1. Standardized, prespecified text to be read by study participants to collect voice recordings.**

Universal Declaration of Human Rights, United Nations.

**English**

Everyone has the right to a standard of living adequate for the health and well-being of himself and of his family, including food, clothing, housing and medical care and necessary social services, and the right to security in the event of unemployment, sickness, disability, widowhood, old age or other lack of livelihood in circumstances beyond his control.

**French**

Toute personne a droit à un niveau de vie suffisant pour assurer sa santé, son bien-être et ceux de sa famille, notamment pour l'alimentation, l'habillement, le logement, les soins médicaux ainsi que pour les services sociaux nécessaires ; elle a droit à la sécurité en cas de chômage, de maladie, d'invalidité, de veuvage, de vieillesse ou dans les autres cas de perte de ses moyens de subsistance par suite de circonstances indépendantes de sa volonté.

**German**

Jeder hat das Recht auf einen Lebensstandard, der seine und seiner Familie Gesundheit und Wohl gewährleistet, einschließlich Nahrung, Kleidung, Wohnung, ärztliche Versorgung und notwendige soziale Leistungen gewährleistet sowie das Recht auf Sicherheit im Falle von Arbeitslosigkeit, Krankheit, Invalidität oder Verwitwung, im Alter sowie bei anderweitigem Verlust seiner Unterhaltsmittel durch unverschuldete Umstände.

**Portuguese**

Toda a pessoa tem direito a um nível de vida suficiente para lhe assegurar e à sua família a saúde e o bem-estar, principalmente quanto à alimentação, ao vestuário, ao alojamento, à assistência médica e ainda quanto aos serviços sociais necessários, e tem direito à segurança no desemprego, na doença, na invalidez, na viuvez, na velhice ou noutros casos de perda de meios de subsistência por circunstâncias independentes da sua vontade.
