## Supplementary figures and images for "Development of a vocal biomarker for fatigue monitoring in people with COVID-19"

### SOM2. VGG19 extracted features from participants' audio recordings

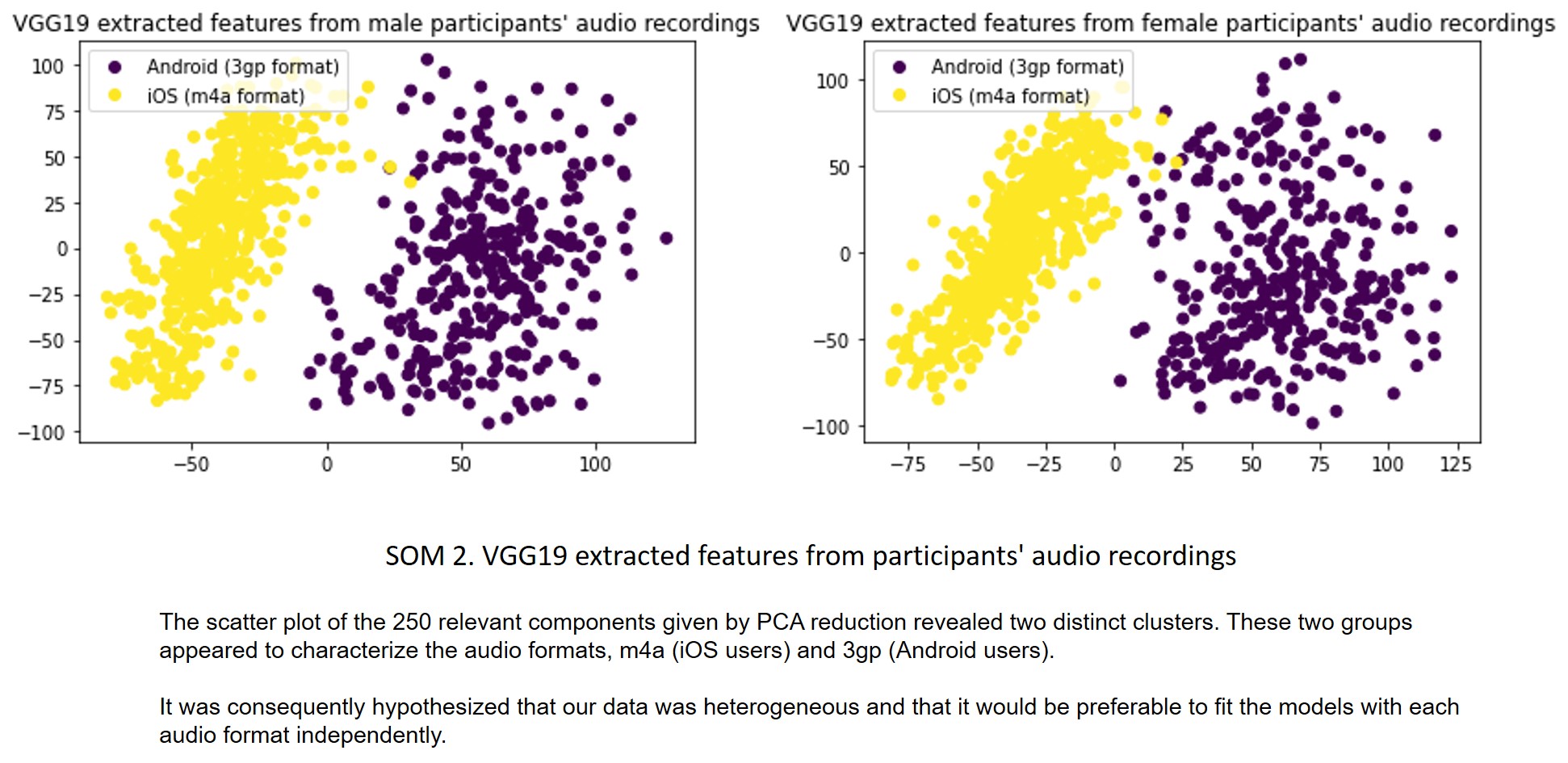
